# Support Experiences Following Unintended Pregnancy in the Netherlands

**DOI:** 10.64898/2026.07.03.26356675

**Authors:** Zeynep Inan, Merel Sprenger, M. Nienke Slagboom, Joyce M. Molenaar

**Affiliations:** Health Campus The Hague/Department of Public Health and Primary Care, Leiden University Medical Center, The Hague, the Netherlands; Population Health and Health Services Research, Center for Public Health, Healthcare and Society, Nationale Institute for Public Health and the Environment (RIVM), Bilthoven, the Netherlands

**Keywords:** unintended, pregnancy, support, parenthood

## Abstract

**Background:** Unintended pregnancies can introduce stress and shift life trajectories. Social support may buffer these effects, yet its influence during an unintended pregnancy and into the early parenthood period is not clear. This study aimed to understand the types and gaps of social support experienced throughout this period.

**Methods:** This study utilized interview data under the RISE UP study in The Hague, the Netherlands. 13 mothers and 8 partners who experienced an unintended pregnancy participated in semi-structured interviews between 2024 and 2025. Interviews were thematically analyzed using House’s social support framework.

**Results:** Different types of support were highlighted across the entire timeline from pregnancy to early parenthood, underlining its dynamic nature. Emotional and instrumental support stood out the most throughout. A key form of emotional support was knowing that support is available, even if not needed immediately.

**Conclusions:** Perceived support during unintended pregnancy is shaped more by contextual factors than by pregnancy intention. While emotional and instrumental support are valued throughout, their form differs by the family’s unique circumstances, emphasizing the need for tailored support across the perinatal and postpartum periods.

## Introduction

Unintended pregnancy is a significant life event that can introduce stress, uncertainty, and shifts in expected life trajectory. Health organisations commonly define unintended pregnancy as a pregnancy that was either *unwanted* or *mistimed* (Auerbach et al., 2023). However, critics of this dichotomous framework suggest that pregnancy intention exists on a continuum, incorporating ambivalence and fluidity in feelings before and after conception (Aiken et al., 2015; Auerbach et al., 2023; Borrero et al., 2015). While unintended pregnancies are frequently associated with adverse outcomes such as maternal depression, pre-term birth, and low birth weight (Nelson et al., 2022), the framing of this experience solely as a risk factor can overlook the dynamic ways in which individuals adapt to stressful life transitions. Firstly, parents may embrace an unintended pregnancy despite having used contraceptive methods (Aiken et al., 2015; Helfferich et al., 2021). Secondly, some people do not feel planning for pregnancy is necessary or even possible (Auerbach et al., 2023; Borrero et al., 2015). Moreover, unintended pregnancy can trigger positive life changes and foster individual agency, especially in contexts where support is available (Golnar Bajegani et al., 2026). The experience of an unintended pregnancy is thus not universal for all but is shaped by contextual, situational, and personal characteristics. A notable characteristic influencing this experience is social support, whose influence is the focus of this paper.

In this study, social support is understood as the experience and perception of being cared for and having available assistance from others, as well as being part of a supportive social network. According to House’s typology (1983), support can take the form of informational, instrumental, emotional, and appraisal support. It can be received from various sources such as one’s partner, family, peers, workplace, or the healthcare system (House, 1983). Social support is thought to promote health by creating a sense of belonging and stability, and by buffering the effects of stress (Cohen & Wills, 1985).

Studies have shown the beneficial effects of social support during pregnancy and in the postpartum period. For instance, mothers with supportive partners expressed low levels of anxiety around mid to late pregnancy (Rini et al., 2006). Limited social support during pregnancy is associated with lower maternal well-being, higher depressive and anxiety symptoms, reduced quality of life, and adverse birth outcomes such as lower weight, shorter length, and preterm birth (Elsenbruch et al., 2007; Rini et al., 2006).

To our knowledge, the relationship between social support and an unintended pregnancy is still underexplored. Kavanaugh et al. (2017) found that social support could buffer the disruptive impacts of an unintended pregnancy, particularly in educational trajectories and financial strain (Kavanaugh et al., 2017). While most parents expressed challenges in these domains, social support could mitigate the adverse effects, with partners or family helping mothers continue schooling by assisting with childcare. However, overall support tended to decline after childbirth (Kavanaugh et al., 2017).

In another study, Newmyer (2024) followed a cohort of young mothers between the ages of 18 and 22 for over two years. The study observed that mothers with unintended pregnancies are more likely to receive support, particularly gifts and conversational support, throughout the pregnancy compared to mothers with intended pregnancies (Newmyer, 2024). In line with Kavanaugh et al. (2017), this study observed that mothers with unintended pregnancies received consistently high support during the first five months of pregnancy, after which the amount of support decreased.

On the other hand, providing social support does not always produce the intended effect: mismatched support can be unhelpful or even harmful. For instance, general research on support has observed that providing more information than the receiver desires can harm relationships (McLaren & High, 2019). On a similar note, Crowley and colleagues (2019) observed that providing high social support when it was not desired as much could lead to diminished affect improvement in unintended pregnancies (Crowley et al., 2019). These studies emphasize the importance of not only the presence of social support but also the right type and amount of support in the proper context.

Past studies have provided some insight into how social support influences various aspects of life following an unintended pregnancy, the amount of support individuals receive, and the importance of matching the support to the recipients’ needs. However, less is known about how individuals perceive different types of support throughout pregnancy and the postpartum period. Furthermore, the existing research has primarily focused on the experiences of only the pregnant person, with relatively little attention paid to the experiences and the support needs of their (former) partner.

To address these gaps, this study draws on in-depth interviews with both the pregnant person and their (former) partner to examine the support parents received during and after an unintended pregnancy, with attention to the mismatch between the support they desired and the support they received.

## Methodology

### Participants

This qualitative study is part of the RISE UP study, a mixed methods study which ran between 2022 and 2025. The RISE UP study included pregnant individuals in the Hague and their (former) partners and aimed to explore the combinations of factors associated with unintended pregnancies in The Hague, the Netherlands.

Eligible participants completed the RISE UP survey, in which the Dutch Adaptation of the London Measure of Unplanned Pregnancy (DA-LMUP) questionnaire measured pregnancy intention (Barrett et al., 2004; Sprenger, Beumer, et al., 2025). DA-LMUP scores of 9 and below are classified as an unintended pregnancy as advised by LMUP guidelines (Barrett et al., 2004; Sprenger, Beumer, et al., 2025). Participants who reported an unintended pregnancy were invited for two interviews, one during pregnancy(Sprenger et al., 2026). and one when the baby was approximately six months old. This paper focuses on the second interview on support experiences.

### Qualitative Research: Data Collection and Analysis

Pregnant people and their partners were recruited through convenience sampling at various sites such as midwifery clinics, Youth and Family Centres, GP practices, and social media advertisements. To minimize selection bias, additional recruitment efforts were directed toward locations with lower participation rates. A detailed description of the recruitment procedures has been published previously by Sprenger et al. (2025).

Fourteen interviews were conducted by MS as part of the RISE UP study, comprising 7 with both parents, 6 with just the mothers, and 1 with just the father. Participants had an average age of 31 years, with ages ranging from 20 to 40. The topic guide for this interview can be found in the RISE UP study project on the Open Science Framework: https://doi.org/10.17605/OSF.IO/63HBE. The interviews were conducted in Dutch, English, or a mix of the two. All interviews, which lasted between 33 and 145 minutes (median 71 minutes), were transcribed verbatim. ZI thematically coded and analyzed the interviews using Atlas.ti version 24.02.0. The first three interviews were coded in collaboration with JM, and the analysis was discussed in weekly meetings within the supervisory team to strengthen credibility and interpretation.

The coding process was guided by House’s typology of social support, which distinguishes between emotional, instrumental, informational, and appraisal support (House, 1983). Emotional support refers to acts that provide love, trust, empathy, and caring. Instrumental support encompasses tangible behavior that directly helps the person in need. This can take the form of labour, money, or manipulating the environment. Informational support refers to any information provided that allows the recipient to deal with challenges. Lastly, appraisal support entails information that the recipient can use to self-evaluate, such as affirmations and feedback (House, 1983). The coding scheme was initially constructed deductively, using House’s typology, time period, and source of support. Then, inductive coding was incorporated to remain close to the original material.

The results are organized temporally, beginning with the pregnancy period, followed by the birth event, the immediate postpartum period, and early parenthood. The immediate postpartum period begins directly after birth and encompasses the postnatal care period (*kraamzorg*) and the initial weeks of living with the baby. Early parenthood refers to the period following the immediate postpartum phase up to the time of the interview, when the baby was approximately six months old.

## Results

Findings on the types of social support that parents – thirteen mothers and eight fathers – received are summarized in Table 1 and described in more detail in the text. Findings on support gaps and mismatches are described in the text. All findings are organized by the timeline, which begins with pregnancy and ends with the early parenthood period.

**Table 1.** Support Types and Sources across Timeline.

| *Type / Time* | *Pregnancy* | *Birth* | *Immediate Post-Partum* | *Parenthood* |
| --- | --- | --- | --- | --- |
| *Emotional Support* | • Midwife: personal attention<br>• Friends, family and partner: listening ear<br>• Friends and family: the sentiment that support is available if needed<br>• Friends and family: being happy and enthusiastic for the pregnancy.<br>• Friends: accompanying to appointments<br>• Friends: affirming difficulty of the situation | • Healthcare Workers (HCW): calm demeanor<br>• HCW: checking up on mothers wellbeing<br>• HCW: shared decision making<br>• Friends, partner and HCW: sticking to the birthplan<br>• Friends and partner: presence | • Maternity care assistant (MCA): listening to parent's wishes<br>• MCA: asking how mother is doing.<br>• Friends, family: visiting | • Friends, family and partner: listening ear<br>• Friends with children: complaining together, feeling understood.<br>• Friends and family: the sentiment that support is available if needed<br>• Friends and family: going on walks together, helping the mother get outside of the house, distraction<br>• Partners: mental support |
| *Instrumental Support* | • Friends, family and partner: helping with chores<br>• Friends and family: cooking or bringing food<br>• Family: caring for other children | • Family: caring for other children | • MCA: providing food, household chores<br>• MCA: infant and childcare<br>• Friends and family: providing food, practical gifts<br>• Friends and family: handles chores<br>• Family: caring for older children. | • Family, childcare services, babysitters: childcare<br>• Work: flexibility and reduced load, distraction |
| *Informational Support* | • Baby courses: useful information | • HCW: guidance during birth | • MCA: infant care explanation<br>• Breastfeeding instructor: information, guidance | • Children's Health Clinic: information on breastfeeding and sleep schedule.<br>• Parent friends: childrearing tips |
| *Appraisal Support (Esteem support)* |  | • MW: having confidence in mother |  | • Parent friends: confirmation that 'things are going well' |

### Throughout the Timeline

Some forms of support were appreciated throughout the timeline, rather than being limited to a specific period. The sentiment that support is available if needed was highly valued by parents at all times:

> *“For me, connection is the most important thing. I’ve noticed this both in parenthood and during pregnancy. The feeling that you have people around you and that you can connect with them, if necessary, and that they are there.” (Mother, 36–40 years)*
>
> *“When we ask for help, there are always people there for us.” (Father, 31–35 years)*

For instance, multiple parents appreciated that their family was always present and ready to help if they had appointments and needed someone to care for the older child. Emotional support, in the form of listening, was prominent during the pregnancy stage but also appreciated in every phase:

> *“…And friends, to complain occasionally. A listening ear. I think that’s about it. And the same goes for after childbirth.” (Mother, 31–35 years)*

Instrumental support from friends and family in the form of providing food, assisting with childcare and household chores, and bringing practical gifts was appreciated in all periods.

Notably, the unintendedness of the pregnancy did not surface in recounts of support experiences organically. Only when asked to reflect on their unintended pregnancy, two single mothers described missing the emotional support of having the child’s father, or a partner, actively involved and emotionally invested in the pregnancy and their lives. These mothers described loneliness:

> *“There’s just no one else in the world who gets that excited about your child picking something up for the first time. You have to go through those moments much more on your own, and that seems like one of the hardest parts of single parenthood to me.” (Mother, 31–35 years)*

### Pregnancy

During pregnancy, parents primarily mentioned receiving emotional and informational support. As the main form of emotional support, many parents appreciated having a ‘listening ear’. This meant having someone to talk to, complain to, and get reassurance from. Partners, friends, or family members provided this type of support. However, the source of this support could be important, as one single mother explains:

> *“Well, I noticed during pregnancy that I missed having a partner I could complain to. I did that with friends from time to time, but you notice it’s different because people also have their own lives and are busy. It’s different when you’re with someone and you can vent for a while.” (Mother, 36–40 years)*

Many parents also described reactions to the pregnancy as a form of support. Parents appreciated that family and peers expressed excitement about the pregnancy:

> *“Everyone was very happy for me and really sympathised with me. I think that’s what I got the most out of and needed the most. Because practically, it was tough, but it was okay… I could do everything myself, like I was already doing.” (Mother, 36–40 years)*

In two cases, participants particularly valued support from social circles that acknowledged the unintended nature of the pregnancy and the mixed emotions it could evoke. For example, one mother described feeling dismissed when her negative emotions about the pregnancy were met with an overly optimistic response, leaving her feeling unheard.

Most parents used baby courses for informational support. Baby courses were evaluated as rich in information, especially for first-time parents. However, some parents found the information outdated, contradictory, or overly detailed. One mother attended these courses not for the informational content but for socialising. In this sense, the baby courses could also serve as a source of emotional support.

The midwife had a central role in providing both emotional and informational support. Midwives made mothers feel taken seriously by listening to them, addressing their concerns, and preparing them for the challenges ahead. Midwives also instilled confidence in the mother that they could handle the birth and what comes after, which could be considered a form of appraisal support. On the other hand, while mothers who were said to have a ‘medical’ pregnancy were satisfied with the medical care they received in the hospital, they missed the personal relationship in midwifery care. One mother even requested that her care be returned to the midwife:

> *“Suppose another child is coming, […] even if you call me medical, I will go to the midwife during my pregnancy. It’s because you are guided rather than controlled. You know those ladies by their first names. They know you by first name. They also ask, ‘Hey, how was it?’ Last time you mentioned this, but what about that? This is so much more personal.” (Mother, 31–35 years)*

### Birth

During birth, parents primarily reported receiving emotional support, which provided a sense of safety. The partner, close friends, and the midwife could provide this support for example by ensuring the hospital team followed the birth plan. Even the mere presence of a partner or close friends during birth helped create a safe environment:

> *“I really liked that they were there; that made me feel safe. I knew that if something wasn’t going well, they would take that up. It wasn’t necessary, but that you know they’re there, that’s nice.” (Mother, 36–40 years).*

Other healthcare workers, such as nurses and gynaecologists, were also an important source of emotional support. Many mothers appreciated the calm demeanor of healthcare workers, which in turn made them feel reassured that things were going alright. One mother humorously described:

> *“One nurse had a reassuring voice, and very calm too. I also had morphine, so I said in between: I’m so glad you talk so calmly, that is so good! That was so funny. But I was happy about it, I really felt supported by them.” (Mother, 36–40 years)*

Guidance and shared decision-making combined emotional and informational support. Mothers appreciated guidance from healthcare workers during the birth process, including when to push and preferred positions. It was also essential to be taken seriously, for instance, during decision-making about getting an epidural.

Instrumental support was primarily mentioned by parents with multiple children. In these cases, grandparents would help by picking up or caring for the older children. Informational support was rarely discussed in this phase. Appraisal support briefly surfaced when a mother expressed gratitude for the midwife who had confidence in her during labour. She described how the midwife’s relaxed demeanour, even doing a puzzle book during labour, reassured her:

> *“My midwife had a lot of confidence in me, and that makes it very pleasant. I also liked that she was working on that puzzle book, for example. It’s fantastic. It exudes a sense of: it’s going as it should. This is totally fine. You don’t have to be afraid of anything.” (Mother, 31–35 years)*

Father’s experiences of support during birth were often less mentioned. Fathers appreciated being able to be in the delivery room during childbirth, which was an improvement for some from their previous experience with their first child.

### Immediate post-partum

This phase covers approximately the first week after birth, starting immediately after birth. Instrumental support in many forms was received and appreciated in this phase.

The most prominent source of support was the maternity care assistant (MCA)^1^. First-time parents appreciated guidance on newborn care, receiving valuable tips and practical information. Parents with multiple children, on the other hand, appreciated the instrumental help from the MCA in the form of childcare and assistance with household chores. MCA support was especially appreciated when the family lived far away or if a parent returned to work. Parents preferred experienced and self-confident MCAs, who would get things done without being asked. However, some felt the help was too lengthy and wanted to spend time alone with their family.

On the other hand, three sets of parents faced a support gap because they missed out on receiving additional MCA assistance after a complicated birth, as they were no longer eligible following their extended hospital stay. While they were satisfied with the hospital care, they felt they did not learn how to care for the baby:

> *“[At the hospital] it was survival. I think that when you are just at home with the maternity care assistant, you also learn a lot. [At the hospital], it was kind of a survival mode until you are better enough to go home, and we hadn’t learned anything.” (Mother, 26–30 years)*

### Early Parenthood

During this period, parents received considerable instrumental support in the form of childcare. This helped by allowing parents to get out of the house, pursue social activities, and get some rest. Similar to the immediate post-partum period, this form of support could be limited by the family living far.

> *“My parents were very supportive with all the appointments and things like that, and with looking after [child 1]. They never made it seem like a burden. They were truly there for us.” (Father, 31–35 years)*

Informational support was also mentioned and appreciated in this phase, particularly from Child and Youth Healthcare service (CYH) workers, whom they turned to for questions about their children. Most parents characterized CYH workers as kind and helpful. However, some, especially those with experience or in related healthcare fields, felt the information was outdated, inconsistent, or invalid.

Parents appreciated having friends or other acquaintances who are also parents or are in the same stage of life. This meant they could chat about their shared struggles and joys, or receive appraisal support, improving their confidence in their abilities as parents:

> *“I have a good colleague. She also lived nearby for a while at the time. I was able to chat with her a couple of times. Just to confirm, it’s actually going really well. “(Mother, 26–30 years)*

Some parents also mentioned that they felt lonelier during this period and that their friendships had changed. This was usually the case when the friends did not have children themselves:

> *“Friendships change. I notice that people are not so good at assessing the impact of having a child, and that they expect you to continue at the same pace, the same way of life.” (Mother, 26–30 years)*

This loneliness was also evident when families lived far away or when friends’ children were in a different age group. Some mothers who lacked friends tried to build a new social network through activities centered on motherhood. Fathers experienced more difficulty, noting that most pregnancy-related social activities target mothers, and men have fewer spaces to connect in their role as fathers:

> *“I think there must be a way of doing more with dads. [….] Women with young children see each other and go ‘Let’s go and get a coffee sometime’ and then they go and get a coffee together sometime and it’s nice. And dads go ‘Alright’ and that’s it.” (Father, 36–40 years)*

Notably, many parents described a shift in both the need for and availability of support during this period, often noting that they mainly managed on their own. Some parents highlighted that they leaned more on their partners and the feeling of being a team during this period. As one father noted,

> *“It’s much easier to carry twice as much, twice as many things if I’m only carrying half of everything, rather than carrying the full weight.” (Father, 26–30 years)*

## Discussion

This study aimed to explore how parents of unintended pregnancies perceived and experienced the social support they received during and after the pregnancy. Findings showed that both desired and received support were dynamic, varying across pregnancy, birth, the postpartum period, and early parenthood. Insights into support gaps and mismatches were limited to distance to extended family and loneliness of single mothers, as parents were mainly satisfied with the support they received.

Emotional and informational support were particularly valued during pregnancy and birth. This aligns with findings from the meta-analysis by Al-Mutahwah et al. (2023), which emphasized the importance of emotional support during this period. The review also noted that informational support can function as emotional support by helping mothers feel cared for and reassured (Al-Mutawtah et al., 2023; Chongo & Ngoma, 2014). After the immediate postpartum period, many parents mentioned needing and receiving less support, consistent with Kavanaugh (2017). Parents rarely mentioned appraisal support. This may indicate that appraisal support played a limited role in their experiences, although it is also possible that participants did not recognize such interactions as a distinct form of support.

Crucially, parents emphasized the importance of feeling that a support network is present and ready to help, even if not needed immediately. The perceived availability of support appeared to matter as much as, or even more than, the amount of actual support received. This supports the direct-effect hypothesis of Cohen and Willis (1985), which states that believing in the availability of social support promotes health by providing a sense of belonging and stability. Similar findings have been reported in perinatal research, where perceived social support is associated with better well-being (Elsenbruch et al., 2007; Rini et al., 2006).

While findings suggest that most parents were satisfied with the support they received, parents whose families lived abroad or across the country seemed to face a significant support gap. Participants with distant families often missed instrumental support during the immediate postpartum and early parenthood periods, as the distance limited their families from assisting with practical matters, such as chores, childcare, and food provision. Nevertheless, many of these participants could engage alternative social support networks through friends, partners, and local initiatives. These insights emphasize the need for tailored support for international families or those without close access to their extended family networks.

This study has several strengths. The interviews explored experiences across multiple phases of the pregnancy and postpartum period, providing insight into how support needs and experiences evolved over time. Additionally, the inclusion of both mothers and partners provided a broader perspective on support dynamics. The application of House’s social support framework further strengthened the study both conceptually and methodologically.

However, limitations should be considered when interpreting the findings of the paper. The interview guide primarily focused on types and sources of support rather than on participants’ lived experiences of receiving support. This may have constrained the insight into how and why certain forms of support were perceived as helpful or unhelpful across different stages of the perinatal period.

## Conclusions

These findings highlight the need to recognize the dynamic nature of pregnancy experiences and social support. Participants primarily described support in relation to their changing circumstances, needs, and relationships over different time points during pregnancy and the postpartum period. On the other hand, the initial intendedness of the pregnancy was rarely mentioned as influencing their experiences. Parents valued emotional and practical support, especially knowing support was available. For health and social care during pregnancy and after, the findings suggest proactively assessing support needs and mismatches at a given moment, rather than just early in pregnancy. Future research should explore what makes support feel helpful in different phases. More understanding is needed surrounding how parents experience support and evaluate the helpfulness of specific support. These insights could help shape care that better matches what people actually benefit from in different situations.

## List of abbreviations

CYH: Child and Youth Healthcare services
HCW: healthcare worker(s) including midwife, doctors, and nurses
LMUP: London Measure of Unplanned Pregnancy
MCA: maternity care assistant
MW: midwife
RISE UP: RISk and rEsilience in Unintended Pregnancy

## Declarations

### Ethics approval and consent to participate

The RISE UP study has been granted non-WMO ethics approval under code N21.127 by the Medical Research Ethics Committee (METC) of Leiden-Den Haag-Delft. All participants are informed of the study aim, participation risks, and benefits through an information letter and follow-up questions. They are then asked to fill out an informed consent form. Interviews were anonymised during transcription. Data storage and access are in line with the GDPR.

### Consent for publication

Not applicable

### Availability of data and materials

Interview data is personal and confidential and therefore not available.

### CRediT Roles

Zeynep İnan: Data curation, Formal analysis, Writing - original draft, Writing - review & editing.

Merel Sprenger: Conceptualization, Methodology, Investigation, Data curation, Supervision, Writing - review & editing.

M. Nienke Slagboom: Conceptualization, Methodology, Supervision, Writing - review & editing.

Joyce M. Molenaar: Formal analysis, Supervision, Writing - review & editing.

### Declaration of generative AI use

Generative AI was used solely to improve the flow of text.

## Acknowledgements

We would like to thank all parents who participated in this study and generously shared their stories and experiences. We also thank the individuals and organizations who assisted with participant recruitment. This study would not have been possible without their contributions.

## Footnotes

1 Maternity care assistant (*kraamverzorgster* in Dutch) is a type of medical service worker unique to the Netherlands. This service provides postnatal care during the 8-10 days after birth. The maternity care assistant checks up on the baby’s and mother’s health status, assists in breastfeeding, provides information on baby care and help with the care of other children of the household. They might also help with household chores. This service is mostly covered by the post-natal care insurance package, with some co-payment(Ministerie van Volksgezondheid, 2017).

